# Medical students in India favor reforms in teaching-learning, clinical training, and evaluation methods

**DOI:** 10.1101/2025.07.16.25331640

**Authors:** Abhinav Jha, Paras Goyal, Aryan Erry, Prachi Renjhen, Ravi Prakash Jha, Aparna Gupta, Utsav Rajvanshi, Saran Singh

## Abstract

**Background:** Despite the implementation of Competency-Based Medical Education (CBME) in India, there is limited data on the perspectives of students regarding prevailing educational methodologies.

**Methods:** An observational cross-sectional study was conducted to assess the perception and satisfaction of Indian medical students towards current teaching-learning and assessment methods. Data were collected via an electronic questionnaire, stratified by academic year, and analysed using SPSS.

**Results:** (N=413) 45% students expressed a neutral response to the curriculum, with greater satisfaction in the clinical years (44.9%) than pre/para-clinical years (30.7%). Lectures were rated neutral by 53%. Tools like clinical demonstration videos (68.8%) and animated videos (56.7%) were favoured over traditional methods. Outpatient and bedside learning were the preferred practical instruction modes (58.9%), though poor departmental coordination was cited as a significant barrier to clinical learning by 42.5%. Students favoured MCQ-based assessments, complemented by viva-cum-practical (78% in clinical years, 68.8% in pre/para-clinical years), while written long essay-type questions were less preferred.

**Conclusions:** Students prefer interactive, clinically integrated teaching and MCQ-based assessments. However, as of now, preferences of students seem to differ from reality, especially in developing countries like India. Addressing these concerns and feedback from students can guide in-depth discussions for medical education curriculum reforms in resource-limited settings.

## Introduction

In India, the Graduate Medical Education (GME) Regulations and National Medical Commission (NMC) aim to develop Indian Medical Graduates (IMGs) equipped with the essential knowledge, skills and competencies to serve as the first point of contact in the healthcare system^1^, with the overarching goal of producing competent and effective physicians. To meet this goal, curriculum evaluation becomes critical.^2^ The curriculum comprises the ‘what, why, how, and how well’ of student learning and is implemented through a combination of teaching-learning and assessment methods.^3^

Curriculum effectiveness depends on the alignment of teaching-learning and assessment methods. Medical teaching involves both knowledge dissemination by the teacher, and its application by students.^4^

In a medical school, teaching-learning methods are different across pre-clinical, para-clinical and clinical phases of training. Teaching-learning methods commonly used are-didactic lectures, seminars, small group teaching or discussions, practical teaching, self-directed learning (SDL), bedside clinics, demonstrations, skill labs, and various others. Modernized methods available are-case-based, problem-based, simulation-based, audience response systems (ARS), social media and video lectures (e-learning), peer-assisted, flipped classroom, etc.^5^

The efficacy of healthcare delivery is inarguably dependent on the knowledge and skills of doctors. Assessments play a crucial role in consolidating both knowledge and skills. If we look deeper into this, the quality of “methods of assessment” are one of the key factors of the long-term memory and skills which students develop.^6^ Assessment methods used mostly are– theoretical written (long answer Type, short answer type, multiple choice questions (MCQ), matching items, etc.), viva voce, logbooks, skills assessment and Objective Structured Clinical Examination (OSCE)^5^.

Different teaching-learning and assessment methods greatly affects the outcome of future performance and intellectual ability to manage patients as doctors. There is a lack of recent data regarding perception of medical students towards currently operating teaching-learning and assessment methods in Indian medical schools. This study aims to assess the perception of medical students towards the currently operating teaching-learning and assessment methods in an Indian medical school, and to identify particular barriers to effective instruction.

## Materials and Methods

An observational cross-sectional study was conducted over a two-month period among medical students at a medical school in New Delhi, India. Ethical approval was obtained from the Institutional Ethical Committee (IEC). Informed electronic consent was obtained from all participants, and anonymity was maintained throughout the study.

A pilot study was conducted to estimate the required sample size. Using the standard sample size formula 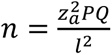 (95% confidence interval, 50 % response distribution, 0.05 margin of error), the minimum sample size was calculated to be 384 participants.

Data collection was carried out through a structured electronic questionnaire created using Google Forms, which included an embedded consent form and participant information sheet. It was disseminated through official batch-wise WhatsApp groups. Two versions of the questionnaire were developed: one for pre- and para-clinical year students (first- and second-year), and another for clinical-year students (third- and fourth-year), to ensure content relevance.

Both questionnaires consisted of three sections. Section 1 was common to both groups and included seven questions assessing students’ overall perception of the curriculum and theory lectures, including one question on a 5-point Likert scale evaluating satisfaction with various teaching-learning aspects.

Section 2 was group-specific. Pre- and para-clinical year students responded to one Likert-scale question focused on practical/laboratory sessions. Clinical-year students answered seven questions examining their satisfaction with clinical postings, perceiving barriers to effective learning, and preferences for basic life support (BLS) and basic surgical skills training.

Section 3, common to both groups, consisted of seven questions assessing students’ perceptions and preferences regarding various modes of assessment. One question required students to select multiple preferred modes of instruction. While response options were largely consistent across both versions, the wording related to MCQ-based assessments differed slightly to reflect group-specific academic contexts. Pre- and para-clinical students were presented with a combined MCQ/viva/practical option, whereas clinical-year students selected separately between MCQs and viva/practical assessments. This adaptation ensured contextual relevance while maintaining data comparability.

Statistical analysis was performed using IBM SPSS Statistics Version 30.0. Associations between categorical variables (such as year of study or gender and teaching-learning or assessment preferences) were analysed using Chi-squared tests. A p-value less than 0.05 was considered significant.

## Results

**Table 1:** Participant Demographics and Academic Distribution (N=413)

| Variable (N=413) | Percent (n) |
| --- | --- |
| **Academic Professional Year** |  |
| MBBS Phase 1 (1<sup>st</sup> Year) | 29.1 % (120) |
| MBBS Phase 2 (2<sup>nd</sup> Year) | 19.1 % (79) |
| MBBS Phase 3 Part 1 (3<sup>rd</sup> Year) | 23.0 % (95) |
| MBBS Phase 3 Part 2 (4<sup>th</sup> Year) | 28.8 % (119) |
| **Sex** |  |
| Male | 79.9 % (330) |
| Female | 20.1 % (83) |

A total of 413 medical students participated in the study, with the highest participation from first-year (29.1%) and final-year (28.8%) students. Males constituted 80% of the sample, indicating a significant gender disparity. (Table 1)

**Table 2:** Preferences for Clinical Teaching Methods Among Medical Students (N = 214)*

| Clinical Teaching Method | Students Preferring (%) | Number of Students (n) |
| --- | --- | --- |
| Bedside/OPD based learning | 58.9 % | 126 |
| Theory teaching followed by case presentation | 52.3 % | 112 |
| Long case presentation with discussion | 50.9 % | 109 |
| Multiple short cases with positive finding | 50.5 % | 108 |
| Hands on skills learning | 37.9 % | 81 |

\*Percentages may not total 100% as students could select more than one preferred method

Among the clinical teaching methods, bedside or OPD-based learning was the most preferred (58.9%). Integrated teaching that combined theoretical instruction with case presentations was selected by 52.3 % of students. Multiple short case discussions (50.5%) and long case presentations (50.9%) were nearly equally favoured, while 37.9% preferred hands-on skill learning. (Table 2)

**Table 3:** Comparative Analysis of Curriculum Satisfaction Across Gender and Academic Phase (N=413)

| Variable<br>(N=413) | Percent (n)<br>Total | Percent (n)<br>Males | Percent (n)<br>Females | Percent (n)<br>Pre- and<br>Para-Clinical<br>Years | Percent (n)<br>Clinical<br>Years |
| --- | --- | --- | --- | --- | --- |
| **Overall MBBS curriculum** |  |  |  |  |  |
| Unsatisfied | 17.0 % (70) | 18.5 % (61)* | 10.8 % (9)* | 18.6 % (37)* | 15.4 % (33)* |
| Neutral | 45.0 % (186) | 42.1 % (139)* | 56.6 % (47)* | 50.8 % (101)* | 39.7 % (85)* |
| Satisfied | 28.0 % (157) | 39.4 % (130)* | 32.5 % (27)* | 30.7 % (61)* | 44.9 % (96)* |
| **Lectures** |  |  |  |  |  |
| Unsatisfied | 16.5 % (68) | 17.6 % (58)* | 12.0 % (10)* | 17.1 % (34) | 15.9 % (34) |
| Neutral | 53.0 % (219) | 49.7 % (164)* | 66.3 % (55)* | 55.3 % (110) | 50.9 % (109) |
| Satisfied | 30.5 % (126) | 32.7 % (108)* | 21.7 % (18)* | 27.6 % (55) | 33.2 % (71) |
| **Practicals/**<br>**Clinical Postings** |  |  |  |  |  |
| Unsatisfied | 11.9 % (49) | 13.3 % (44)* | 5 (6.0 %)* | 15.6 % (31) | 8.4 % (18) |
| Neutral | 39.2 % (162) | 35.2 % (116)* | 55.4 % (46)* | 36.2 % (72) | 42.1 % (90) |
| Satisfied | 48.9 % (202) | 51.5 % (170)* | 38.6 % (32)* | 48.2 % (96) | 49.5 % (106) |
| **Modes of Assessment** |  |  |  |  |  |
| Unsatisfied | 16.9 % (70) | 18.5 % (61) | 10.8 % (9) | 19.1 % (38) | 15.0 % (32) |
| Neutral | 50.4 % (208) | 48.8 % (161) | 56.6 % (47) | 50.3 % (100) | 50.5 % (108) |
| Satisfied | 32.7 % (135) | 32.7 % (108) | 32.5 % (27) | 30.7 % (61) | 34.6 % (74) |
| **Current CBME pattern of testing** |  |  |  |  |  |
| Satisfied | 60.8 % (251) | 61.2 % (202) | 59.0 % (49) | 60.8 % (121) | 60.8 % (130) |
| Unsatisfied | 39.2 % (162) | 38.8 % (128) | 41.0 % (34) | 39.2 % (78) | 39.2 % (84) |

\* p < 0.05

Overall satisfaction with the MBBS curriculum was predominantly neutral (45%), with a higher proportion of satisfied students in the clinical years (44.9%) than pre/para-clinical years (30.7%). Satisfaction with lectures mirrored this pattern, with 33.2% of clinical-year students expressing satisfaction, as opposed to 27.6% in the earlier phases. In contrast, practical and clinical sessions received relatively high satisfaction: 48.2% in pre/para-clinical years and 49.5% in clinical years. Mode of assessment showed moderately positive satisfaction overall (32.7%), but dissatisfaction was notably higher among pre/para-clinical year students. The majority (60.8%) of students across all phases were satisfied with the current CBME pattern of assessment.

**Table 4:** Gender- and Phase-Wise Preferences for Theory Lectures (N=413)

| **Variable** | **Percent (n)**<br>**Total**<br>**(N=413)** | **Percent (n)**<br>**Males**<br>**(N=330)** | **Percent (n)**<br>**Females**<br>**(N=83)** | **Percent (n)**<br>**Pre and Para**<br>**Clinical**<br>**Years**<br>**(N=199)** | **Percent (n)**<br>**Clinical**<br>**Years**<br>**(N=214)** |
| --- | --- | --- | --- | --- | --- |
| **Duration** |  |  |  |  |  |
| 30 minutes | 19.9 % (82) | 20.6 % (68) | 16.9 % (14) | 11.6 % (23)* | 27.9 % (59)* |
| 45 minutes | 58.6 % (242) | 56.7 % (187) | 66.3 % (55) | 61.8 % (123)* | 55.6 % (119)* |
| 60 minutes | 19.4 % (80) | 20.0 % (66) | 16.9 % (14) | 24.1 % (48)* | 15.0 % (32)* |
| 90 minutes | 2.2 % (9) | 2.7 % (9) | 0.0 % (0) | 2.5 % (5)* | 1.9 % (4)* |
| **Timing** |  |  |  |  |  |
| 8-10 am | 56.7 % (234) | 55.8 % (184) | 60.2 % (50) | 75.4 % (150)* | 39.3 % (84)* |
| 2-4 pm | 43.3 % (179) | 44.2 % (146) | 39.8 % (33) | 24.6 % (49)* | 60.7 % (130)* |
| **Duration of**<br>**breaks** |  |  |  |  |  |
| No break | 10.4 % (43) | 11.8 % (39) | 4.8 % (4) | 5.0 % (10)* | 15.4 % (33)* |
| 5 minutes | 25.2 % (104) | 24.8 % (82) | 26.5 % (22) | 28.6 % (57)* | 22.0 % (47)* |
| 10 minutes | 41.6 % (172) | 40.3 % (133) | 47.0 % (39) | 43.7 % (87)* | 39.7 % (85)* |
| 15 minutes | 22.8 % (94) | 23.0 % (76) | 21.7 % (18) | 22.6 % (45)* | 22.9 % (49)* |
| **PPT Sharing** |  |  |  |  |  |
| Shared before the<br>lecture | 74.1 % (306) | 74.2 % (245) | 73.5 % (61) | 69.8 % (139) | 78.0 % (167)* |
| Not shared | 6.1 % (25) | 6.7 % (22) | 3.6 % (3) | 5.5 % (11) | 6.5 % (14)* |
| Shared after the<br>lecture | 19.9 % (82) | 19.1 % (63) | 22.9 % (19) | 24.6 % (49) | 15.4 % (33)* |
| **Faculty**<br>**feedback**<br>**received** | 67.8 % (280) | 67.9 % (224) | 67.5 % (56) | 68.3 % (136) | 67.3 % (144) |

\* p < 0.05

The majority of the students (58.6%) preferred 45-minute lectures, with a slightly higher preference among pre/para-clinical year students (61.8%) compared to clinical-year students (55.6%). A clear split in lecture timing preferences was noted, with 75.4 % of pre/para-clinical year students preferring morning sessions, while 60.7% of clinical-year students preferred afternoon timings. Ten-minute breaks between lectures were the most favoured (41.6%), and 74.1% preferred lecture PPTs to be shared in advance, especially clinical-year students (78%). 67.8% were satisfied by the feedback they received from the faculty, with no significant differences across gender or phase of training.

**Figure 1:**
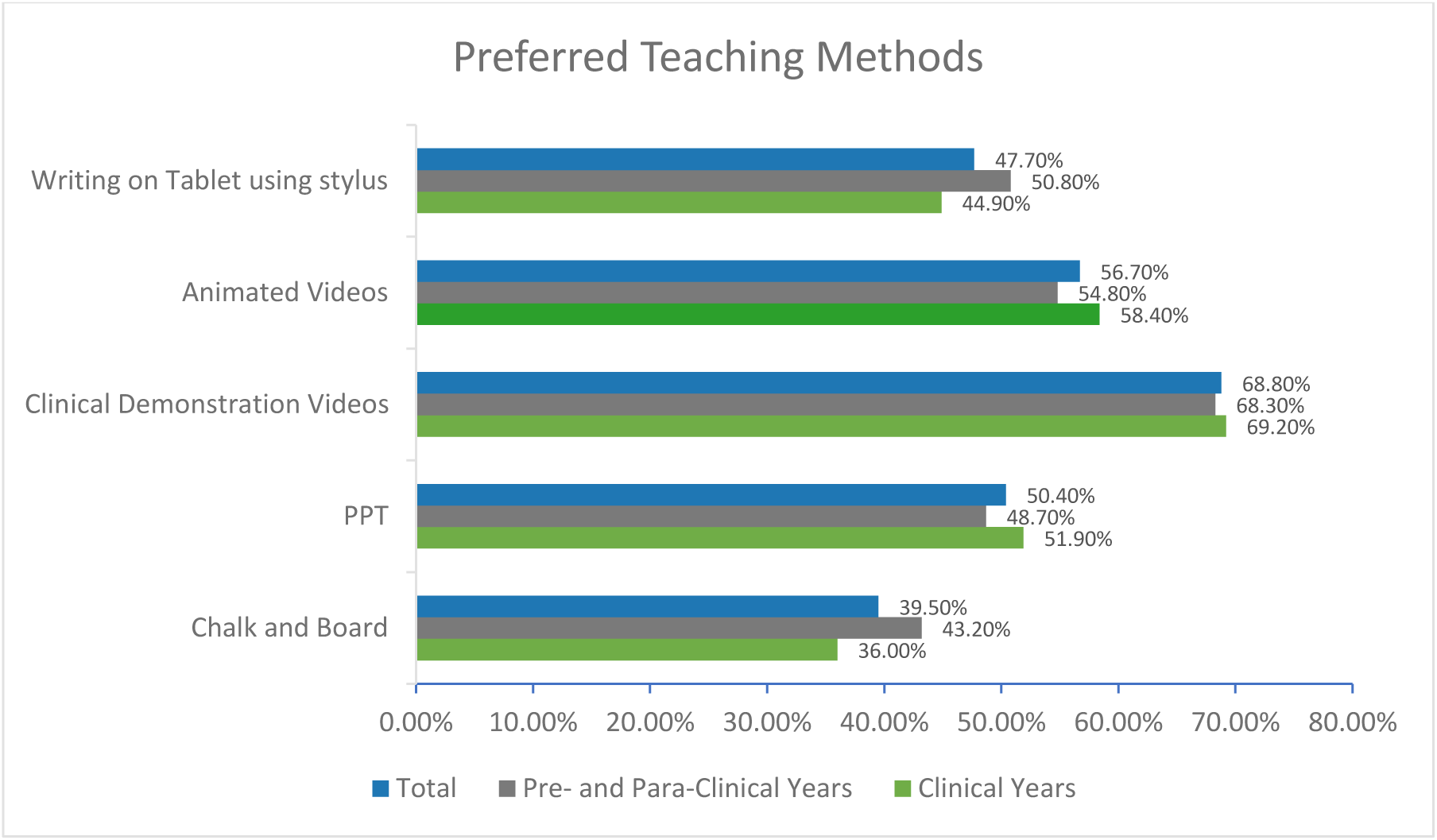
Preferred Teaching Methods

Clinical demonstration videos were the most preferred method (68.8%), followed by animated videos (56.7%) and PPTs (50.4%). Writing with a stylus on tablets was favoured by 47.7% of students.

**Table 5:** Preferred Modalities for BLS and Practical Skills Training Among Clinical-Year Students (N= 214)

| **Variable** | **Percent (n)**<br>**Total** | **Percent (n)**<br>**Males** | **Percent (n)**<br>**Females** |
| --- | --- | --- | --- |
| **<u>Preferred year for BLS and basic surgical skills training</u>** |  |  |  |
| 1<sup>st</sup> Year | 24.8 % (53) | 24.1 % (39) | 26.9 % (14) |
| 2<sup>nd</sup> Year | 28.5 % (61) | 27.2 % (44) | 32.7 % (17) |
| 3<sup>rd</sup> Year Part 1 | 30.4 % (65) | 29.6 % (48) | 32.7 % (17) |
| 3<sup>rd</sup> Year Part 2 | 9.3 % (20) | 11.1 % (18) | 3.8 % (2) |
| Internship | 7.0 % (15) | 8.0 % (13) | 3.8 % (2) |
| **<u>BLS Training Frequency</u>** |  |  |  |
| Every 2 years | 3.7 % (8) | 4.9 % (8) | 0.0 % (0) |
| Every year | 36.9 % (79) | 37.0 % (60) | 36.5 % (19) |
| Every 6 months | 59.3 % (127) | 58.0 % (94) | 63.5 % (33) |
| **<u>Clinical posting Batch Strength</u>** |  |  |  |
| 5 | 8.4 % (18) | 9.9 % (16) | 3.8 % (2) |
| 10 | 41.1 % (88) | 40.1 % (65) | 44.2 % (23) |
| 15 | 38.3 % (82) | 38.3 % (62) | 38.5 % (20) |
| 20 | 12.1 % (26) | 11.7 % (19) | 13.5 % (7) |

Clinical-year students preferred BLS and basic surgical skills training during the 2nd year (28.5%) and 3rd year (30.4%). A majority of them (59.3%) also advocated for biannual training sessions. The ideal batch size was reported to be 10 students (41.1%).

**Figure 2:**
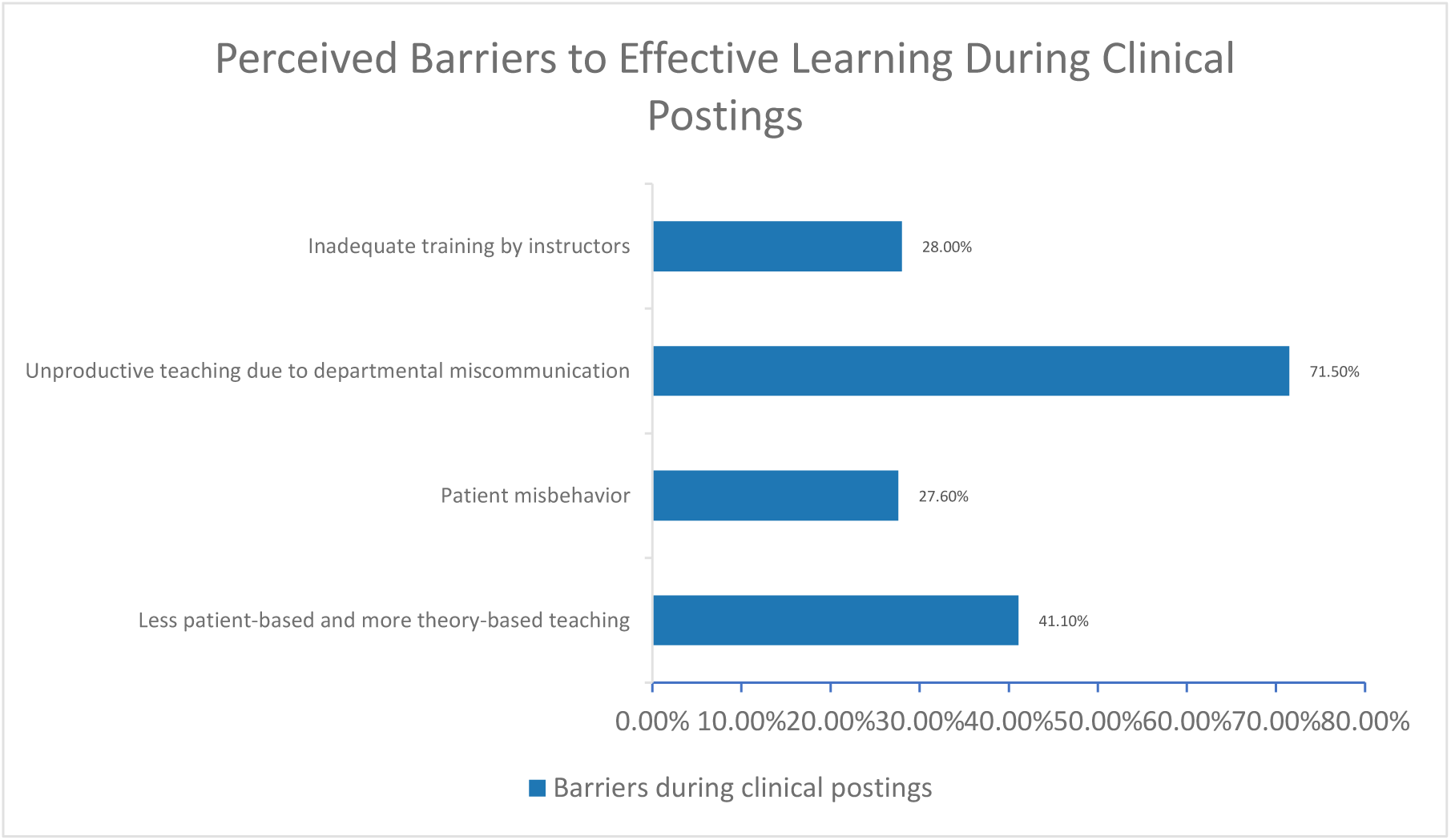
Perceived Barriers to Effective Learning During Clinical Postings

The most cited barrier was poor departmental communication (71.5%). Excessive focus on theory (41.1%), inadequate training by faculty (28%), and patient misbehaviour (27.6%) were other notable concerns.

**Figure 3:**
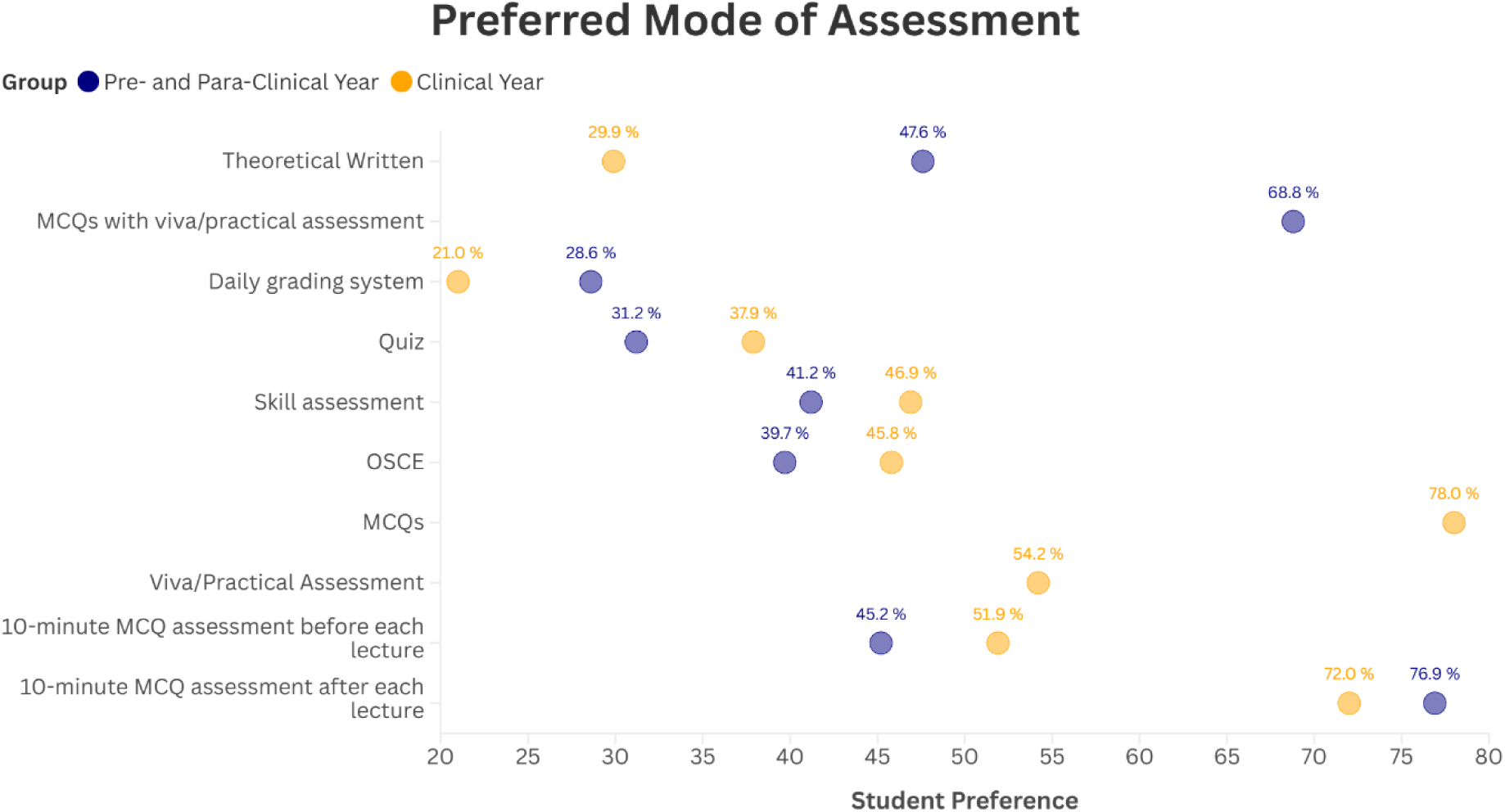
Preferred Mode of Assessment

MCQ-based assessments combined with viva and practical exams were the most preferred format (68.8% in pre/para-clinical, 78% in clinical years). Written theory exams were more favoured by pre/para-clinical students (47.7%) than clinical-year students (29.9%). A substantial 74.1% of students across all years preferred a brief (10-minute) MCQ test at the end of each lecture. Daily grading systems were the least preferred assessment methods.

## Discussion

We found the overall student response revealed a neutral to modestly positive outlook towards the curriculum. Satisfaction levels were notably higher among clinical-year students, likely due to increasing relevance and contextualization of theoretical knowledge as students gain clinical exposure. In contrast, earlier-year students may face a dissonance between their expectations and the didactic nature of pre-clinical education^7^, suggesting a need for early integration of applied learning. There was also a variation in the satisfaction levels with regard to gender, which may reflect differences in learning styles and preferences.

The majority of the students expressed a neutral stance toward traditional theoretical lecture methods; however, students strongly favoured 45-minute theory sessions over the standard hour-long classes, with a 10-minute break between lectures. These findings are consistent with previous studies.^8^ PowerPoint presentations supplemented with clinical demonstrations and animated videos were among the preferred teaching tools, reflecting a demand for interactive tools and multimodal approaches that may help sustain student attention and comprehension. Similar trends have been reported in earlier research^9^, where a significant proportion of students favoured practical demonstrations during lectures, as they enhanced subject understanding, and opposed longer class durations. This indicates a broader shift in student preferences over the past decade, with recent studies supporting technology-assisted and engaging teaching methods, in contrast to earlier preferences for chalkboard-based instruction.^10^ Students also preferred that professors share lecture presentations in advance, likely to facilitate note-taking and allow preliminary exploration of the subject. This preference emphasizes the need for structured and student-friendly delivery.

Differences were noted in lecture timing preference, with clinical-year students favouring afternoon lectures and pre/para-clinical year students preferring morning lectures. This contrast may reflect adaptation to existing schedules rather than inherent preference, potentially influenced by status quo bias. Status quo bias is defined as a cognitive bias that describes the irrational preference for an option only because it preserves the current state of affairs.^11^ Our findings contrast with Bommeri et al’s findings, who found that students find early morning lectures inconvenient.^12^

In contrast, students reported higher satisfaction with practical learning. Outpatient and bedside teaching were the most valued modes of instruction, emphasizing the importance of experiential, patient-centred learning. These preferences echo previous literature, demonstrating that such formats promote the development of clinical skills, foster communication skills, and effectively integrate theory with practical lessons.^13^

During clinical postings, students expressed nearly equal preference for discussing multiple short cases or a single long case discussion, conducted in small groups, ideally fewer than 10 students. This reinforces the value of close faculty-student interaction and interactive teaching. Preference for smaller group sizes has been similarly reported in other studies. Evidence suggests that students reporting undesirable clinical experiences were more likely to feel that large groups interfered with learning. Moreover, the probability of a student having a desirable clinical experience was much higher in groups of 2 students rather than 4.^14^ Although such group sizes would be ideal, implementation may be impeded by resource constraints and poor teacher-student ratios. In many Indian medical schools, the group size for clinical postings may exceed 30 to 40 students per instructor, which can hinder effective clinical teaching. Education policymakers should consider reducing the maximum number of students during clinical rounds to better align with learner preferences, ideally targeting groups of 10 or fewer. Small group discussions have become increasingly supported in literature, as it promotes development of critical thinking and clinical reasoning, skills that are essential for physicians.^15^ Case-based learning has also been consistently identified as a preferred method in recent literature.^16^

A common concern raised by clinical-year students was poor departmental coordination in clinical postings, leading to inefficient utilization of their time. Amaranathan et al. also reported similar results where negative perceptions of clinical teaching were caused by inadequate planning. Often, clinical cases are addressed based on the availability of patients on that particular day in the outpatient clinic, and learning is hindered by overworked physicians and time constraints. Organized and prearranged clinical case-based discussions might help mitigate this issue.^17^ Students expressed a clear preference for initiating Basic Life Support and Surgical Skills Training earlier in the curriculum, ideally during the third year, with reinforcement sessions every six months. This is in contrast to the prevailing norms, where most students learn these skills during internship only. Other studies have shown emphasis upon frequent revisions and constant practice to retain information and abilities in life support. It also allows for updating information or changes in guidelines.^18^

Assessment preferences revealed a strong inclination toward-MCQ based examinations, particularly when supplemented by viva voce and practical evaluations, and these were preferred over written short and long answer questions. This may be influenced by the predominantly MCQ based pattern of postgraduate entrance examinations in India, which is in stark contrast to the subjective, long answer-type and theoretical pattern of undergraduate assessment. Harmonizing formats across levels could ease students’ preparation and improve performance.^19^ Moreover, studies have suggested that MCQ based examinations offer greater ease of preparation, standardization, and efficient testing.^20^ Skills-based assessment and Objective Structured Clinical Examinations (OSCEs) were also widely preferred by students. Previous studies have also shown OSCE to be a widely acceptable assessment tool of knowledge and skills, as it effectively highlights areas of strength and weakness.^21^ Despite these preferences, a majority of students expressed dissatisfaction with current assessment methods. Similar results have also been reported where assessment methods were found to be incomprehensive and inadequate, indicating the need for improvement.^19^ Overall satisfaction with assessment practices remained moderate, underscoring the need for more transparent, diverse and competency-aligned evaluation strategies.

The perceptions of students are clearly shifting toward more interactive, practical and integrated approaches, even though traditional methods retain utility, particularly in the preclinical year. These findings carry important implications for curriculum planners and regulatory bodies. In order to create a system that empowers upcoming physicians with the required knowledge and abilities, it is important for policymakers to reflect student perspectives.

Certain limitations must be acknowledged when interpreting these findings. Although the study had an adequate sample size, responses were self-reported, which could introduce response bias. Additionally, institutional differences may limit generalizability of results, as the study was set in a single centre.

Gender distribution also poses a concern, as statistically significant differences between male and female respondents have been observed in various parameters. Since male participants comprised 80 % of the sample, it is possible that the smaller proportion of female participants could have led to underrepresentation of their views or skewed statistical comparisons. Further research with a more balanced gender representation is needed to confirm these trends.

We also did not control for other potential confounders like socioeconomic status and prior educational background that may shape students’ learning preferences, performance and tendencies, such as burnout.^22^ Multicentric studies across varied institutional settings can help provide more substantial evidence. The study offers useful direction for curriculum reforms in Indian medical education. To foster a learning environment that is both evidence informed and student responsive, ongoing dialogue between the students, faculty, and curriculum development authorities is essential.

## Conclusions

The majority of the students were neutral towards the existing curriculum, with the pre-final and final year students reporting a higher overall satisfaction rate. Theoretical lectures were met with a neutral response, with a preference towards shorter sessions that integrate clinical demonstrations and animated videos alongside traditional PowerPoint based teaching.

The students viewed practical teaching in outpatient and bedside settings as the most suitable method, but cited poor planning and lack of coordination in clinical postings as barriers to effective learning. MCQ based examinations with viva voce and practical evaluations emerged as the most preferred assessment format. These findings suggest a student inclination toward more interactive and effective training and assessment methods. However, as of now students’ preferences seem to differ from the reality especially in developing countries like India. Addressing these concerns and students’ feedbacks can guide in-depth discussions for medical education curriculum reforms in resource-limited settings. The change of pattern inclined towards the preference of students which can be feasible in resource-limited settings can lead to much overall satisfaction of students and hence, may impact long term outcomes as being a doctor. Multicentric longitudinal studies with large, diverse cohorts should be conducted to validate the findings of this study and extrapolate the results. A nuanced understanding of student preferences can assist educational policymakers in implementing meaningful reforms that balance traditional strengths with evolving educational needs.

## Data Availability

All data produced in the present study are available upon reasonable request to the authors.

